# Individual-specific resting-state networks predict language dominance in drug-resistant epilepsy

**DOI:** 10.1101/2025.11.21.25340716

**Authors:** Mervyn Lim Jun Rui, Zhang Shaoshi, Shreya Pande, Xue Aihuiping, Ru Kong, Kareem Zaghloul, Sara Inati, B.T. Thomas Yeo

**Affiliations:** Computational Brain Imaging Group, Yong Loo Lin School of Medicine, National University of Singapore; National Institute of Neurological Disorders and Stroke, National Institute of Health; Division of Neurosurgery, Department of Surgery, National University Hospital

**Keywords:** functional magnetic resonance imaging, precision functional mapping, language lateralization, multi-session hierarchical Bayesian model

## Abstract

**Objective:** To reliably estimate individual-specific resting-state cortical networks and determine if language network topography can predict task-based language dominance in drug-resistant epilepsy.

**Methods:** We utilised a multi-session hierarchical Bayesian model (MS-HBM) trained on drug-resistant epilepsy patients to map high-quality individual-specific cortical networks in this population (N = 65) with only 6 to 24 minutes of resting-state fMRI. We compared the quality of networks to MS-HBM models trained on healthy participants from the human connectome project (N = 40) and tested the generalizability of the model in an independent cohort of drug-resistant epilepsy participants (N = 26). Resting-state language network topography was then used to predict task-based language dominance.

**Results:** Ninety-one participants with drug-resistant epilepsy were included: 61 (67.0%) temporal lobe epilepsy, 29 (31.9%) extra-temporal lobe epilepsy, and 1 (1.1%) undetermined seizure onset zone. The mean age was 33.0 ± 11.4 years and 50 (54.9%) were male. There were 40 healthy participants with a mean age of 29.0 ± 4.0 years, and 16 (40.0%) were male. MS-HBM trained on drug-resistant epilepsy estimated individual-specific networks that more accurately capture cortical functional organization than group-average networks or MS-HBM trained on healthy participants. The trained MS-HBM model generalized to an independent cohort of drug-resistant epilepsy participants with concurrent intracranial electrical stimulation and fMRI. Critically, cortical evoked fMRI activity aligned more closely with individual-specific networks than with group-average networks. Furthermore, individual-specific language network topography significantly predicted task-based language dominance, achieving high accuracy for left (AUC = 0.82), bilateral (AUC = 0.72), and right (AUC = 0.83) dominance.

**Significance:** These results demonstrate that MS-HBM captures functionally meaningful network reorganization in drug-resistant epilepsy and enables accurate, individual-level prediction of language lateralization, with direct implications for presurgical functional mapping.

**Key Points:**

- Existing resting-state fMRI methods have limited ability to predict language dominance for epilepsy surgery.
- Precision functional mapping techniques allow reliable estimation of inter-individual variability in large-scale resting-state networks.
- MS-HBM can map high-quality individual-specific cortical networks in drug-resistant epilepsy using only 6 to 24 minutes of data.
- Individual-specific language network topography predicted task-based language dominance well in individual patients.

## Introduction

Identifying language dominance is a crucial step in planning surgery for epilepsy as it has direct implications for pre-operative counselling and post-surgical language outcomes (Hermann et al. 1994). Current clinical guidelines recommend task-based functional magnetic resonance imaging (fMRI) as a non-invasive alternative to reduce complications from the intracarotid amobarbital test (Binder et al. 1996, Szaflarski et al. 2017, Lado et al. 2024), yet task-based fMRI is highly dependent on patient compliance and task design (Gaillard et al. 2007, Binder et al. 2008). Resting-state fMRI avoids these limitations by removing the need for patient participation, and can map multiple brain functions using a single scan (Fox et al. 2010).

However, prior studies using resting-state fMRI to predict task-based language dominance in drug-resistant epilepsy have reported only modest prediction accuracy (Doucet et al. 2015, Rolinski et al. 2020, Phillips et al. 2021, Pur et al. 2021), possibly because previous methods were unable to fully account for inter-individual variability in language network topography. Furthermore, although numerous studies have reported alterations in resting-state functional connectivity in drug-resistant epilepsy (Bettus et al. 2009, Pittau et al. 2012, Ji et al. 2025, Xie et al. 2025), far less is known about the organization and behavioural relevance of individual-specific cortical networks in this population. Thus, we aimed to characterise individual-specific cortical networks and investigate the clinical utility of the language networks for improving prediction of language dominance in participants with drug-resistant epilepsy using a precision functional mapping approach.

Precision functional mapping is a technique used to delineate functional brain organization using resting-state fMRI at the individual level rather than relying on population-averaged atlases(Laumann et al. 2015, Gordon et al. 2017, Du et al. 2024). This approach is motivated by inter-individual variability in large-scale functional networks (Gordon et al. 2017), as well as systemic differences between healthy and pathological brains (Laumann et al. 2021, Lynch et al. 2024). Thus, mapping the individual’s functional networks in neurological and psychiatric disease (Filippi et al. 2013) holds promise for prognostication (Lynch et al. 2024), network-guided neuromodulation (Kong et al. 2025), and neurosurgical practice (Roland et al. 2019).

A major barrier to clinical translation, however, has been the long scan duration – at least one hour – traditionally required for precision functional mapping (Braga et al. 2017, Laumann et al. 2021, Lynch et al. 2024). We previously developed a multi-session hierarchical Bayesian model (MS-HBM) that overcomes this limitation by enabling reliable estimation of individual-specific networks from limited data: MS-HBM networks obtained using only 10 minutes of data were comparable to other approaches requiring 50 minutes of data (Kong et al. 2019). Individual-specific differences in MS-HBM network topography have been shown to predict inter-individual variation in cognition and mental health (Kong et al. 2019), and aligned with individual-level task-evoked activity (Du et al., 2024). By substantially reducing scan time requirements, this approach might enable precision functional mapping to become feasible in routine clinical settings, where patients might not be able to tolerate long scans.

In this study, we demonstrate that MS-HBM trained on drug-resistant epilepsy patients can be used to map high-quality individual-specific cortical networks in this population using only 6 to 24 minutes of resting-state fMRI. The resulting individual-specific cortical networks conformed better to new resting-state fMRI data from the same participants than both group-average networks and MS-HBM trained on healthy individuals. Furthermore, in an independent drug-resistant epilepsy cohort with concurrent intracranial electrical stimulation and fMRI (Thompson et al., 2020), cortical evoked activity aligned better with individual-specific networks than with group-average networks. Finally, we showed that individual-specific language network topography predicted task-based language dominance well in individual patients. Together, these findings demonstrate that MS-HBM can capture individual-specific network organization in epilepsy and reliably predict language lateralization in individuals.

## Materials and methods

### Participants and Datasets

In this study, we included one open-source dataset of healthy participants and two independent datasets of participants with drug-resistant epilepsy undergoing pre-surgical evaluation. Human Connectome Project (HCP) participants had a mean age of 29.0 ± 4.0 years, 16 (40.0%) were male, and 12 (30.0%) self-reported as non-White and/or Hispanic. NIH epilepsy participants had a mean age of 31.9 ± 11.2 years and 33 (50.8%) were male. Of these, 42 (64.6%) had temporal lobe epilepsy and 23 (35.4%) had extra-temporal lobe epilepsy (Supplementary Table 1). Electrical-stimulation fMRI (esfmri) participants had a mean age of 35.8 ± 11.5 years, and 17 (65.4%) were male. Of these, 19 (73.1%) had temporal lobe epilepsy, 6 (23.1%) had extra-temporal lobe epilepsy, and 1 (3.8%) had undetermined seizure onset zone (Supplementary Table 2).

All datasets included structural and resting-state fMRI scans. In the esfmri dataset, post-operative fMRI was acquired with concurrent intracranial electrical stimulation delivered through stereoencephalography electrodes and interleaved with echo planar imaging volume acquisition (Thompson et al. 2020). Alternating blocks of approximately 30 seconds of no stimulation or stimulation was delivered during the fMRI scan. No cognitive task was given to the participant, and no behavioural effects were evoked during the stimulation period. Pre-processing of imaging data followed the surface-based pipeline of Yeo et al (Yeo et al. 2011, Holmes et al. 2015). Further details regarding the demographics, data collection, and pre-processing are provided in the supplementary methods. After pre-processing, there were 40 HCP participants with 4 runs each. Of the 48 NIH participants with two or more runs, 34 (70.8%) participants were used for the training set, and 14 (29.2%) participants were used for the test set. For the esfmri dataset, there were 11 participants that had at least two valid pre-operative runs and 17 participants that had at least two valid post-operative runs (Figure 1A).

**Figure 1:**
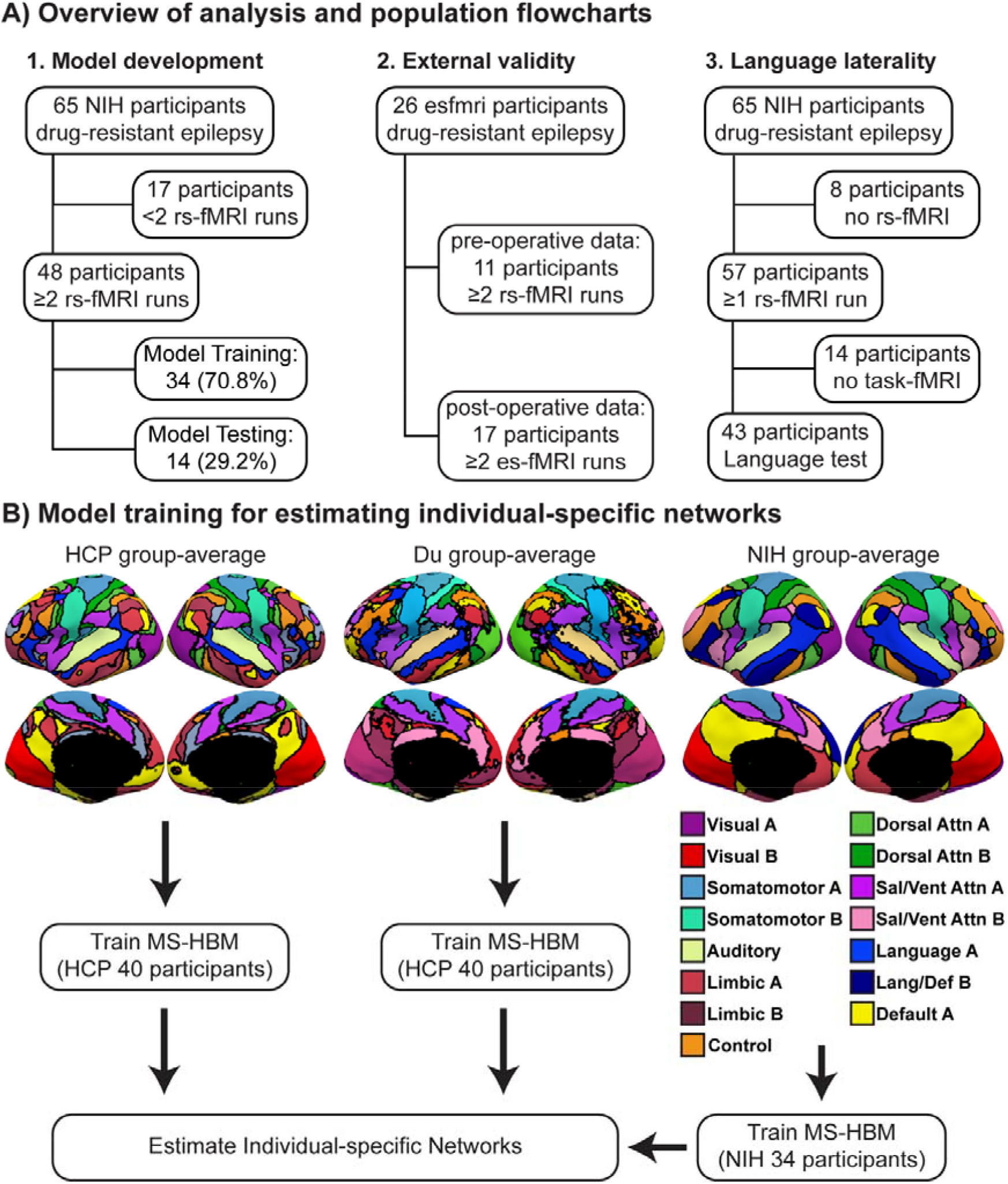
Study overview and population flow chart. (A) We developed a MS-HBM model for estimating individual-specific networks in drug-resistant epilepsy, validated and tested the model in an independent dataset, and showed that the topography of individual-specific language networks was able to predict task-based language dominance. (B) Group-average networks from healthy participants of the HCP dataset, the Du atlas, or drug-resistant epilepsy participants from the NIH dataset were used to train different MS-HBM models. 15-network labels were shown for NIH group-average networks. Networks in drug-resistant epilepsy appeared more focal compared to healthy participants.

### Derivation of group-average networks and training of MS-HBM for estimating individual-specific networks

Our approach has been described previously (Yeo et al. 2011, Kong et al. 2019). At each cortical region (81,924 vertices), Pearson’s correlation between the fMRI time series and 1175 regions of interests were binarized by keeping the top 10% of correlations. The connectivity profiles were then clustered using a mixture of von Mises-Fisher distributions to derive the group-average 15-networks (Lashkari et al. 2010, Yeo et al. 2011). In addition to computing group-average networks for the 40 HCP participants and 34 NIH participants, we also compared our results to group-average networks derived from an independent dataset of 15 intensively-sampled healthy participants (Du et al. 2024). Next, we trained MS-HBM models on either the 40 HCP participants (using the HCP group-average or Du group-average) or 34 NIH participants to estimate model parameters. Further details on the training of MS-HBM may be found in the supplementary methods. Individual-specific networks were visually confirmed using model-free seed-based functional connectivity. Inter-subject similarity in network topography across the 15-networks was quantified using the Dice similarity coefficient across all 57 NIH participants with at least one valid run.

### Validation and generalization of MS-HBM trained on drug-resistant epilepsy

To evaluate the quality of individual-specific networks estimated from MS-HBM trained on drug-resistant epilepsy or healthy participants, we computed two previously established metrics (Gordon et al. 2016, Gordon et al. 2017, Schaefer et al. 2018, Kong et al. 2019): the weighted resting-state connectional homogeneity and weighted task (electrical-stimulation) functional inhomogeneity. These metrics encode the principle that if an individual-specific network captured the system-level organization of the individual’s cortex, each network should exhibit homogeneous connectivity and function.

We first evaluated the resting-state homogeneity of each network on the NIH test set of 14 participants that were not used for training MS-HBM. The generalizability of the MS-HBM model was then tested using pre-operative (11 participants) and post-operative (17 participants) esfmri data, as well as task inhomogeneity during electrical stimulation in the post-operative data. For estimating individual-specific networks in the post-operative data, we used only blocks with no stimulation after discarding the first 4 frames of each block to remove residual effects of the haemodynamic response from the previous block (Pedersen et al. 2021).

To ensure that runs used for estimating individual-specific networks were independent from runs used for computing evaluation metrics, we used a leave-one-run-out cross-validation method. For each participant with n valid runs, individual-specific networks were estimated using n-1 runs, and evaluation metrics were computed on the left-out run. This was repeated for all runs, and an average result was obtained for each participant. Statistical tests were performed using two-tailed paired t-tests between each pair of networks (dof = number of participants – 1). Statistical significance was taken to be q<0.05 after false discovery rate correction.

The average resting-state homogeneity and task inhomogeneity were compared across six networks: (1) HCP group-average networks; (2) individual-specific networks estimated using HCP MS-HBM; (3) Du group-average networks; (4) individual-specific networks estimated using Du MS-HBM; (5) NIH group-average networks; and (6) individual-specific networks estimated using NIH MS-HBM. Resting-state homogeneity was computed by averaging the Pearson’s correlations between each vertex’s time course with the average time course of its network. The correlations were then averaged across all 15 networks while accounting for network size (Gordon et al. 2016, Schaefer et al. 2018, Kong et al. 2019). Task inhomogeneity was computed as the average standard deviation of cortical activation z-scores within each network from unthresholded generalized linear models of intracranial electrical stimulation (Oya et al. 2017). The standard deviation was then averaged across all 15 networks while accounting for network size (Gordon et al. 2016, Schaefer et al. 2018, Kong et al. 2019). A higher standard deviation indicated higher task inhomogeneity. Further details on the computation of the generalized linear models are provided in the supplementary methods.

To visualise the effects of intracranial electrical stimulation on cortical networks, group-average network boundaries and individual-specific network boundaries were overlayed on the electrical-stimulation z-score activation maps. We hypothesised that electrical-stimulation evoked activity would align more closely to individual-specific network boundaries than group-average network boundaries.

### Predicting language dominance using individual-specific network topography

All patients underwent language task fMRI using an auditory description decision task as described previously (Gaillard et al. 2007, Rolinski et al. 2020). Task-based language dominance was determined by a trained epileptologist (SI) through visual inspection of activation maps in the frontal and temporal regions at three thresholds (p < .01, p < .001, top 10% of activations) and confirmed at a multi-disciplinary epilepsy conference. There were a total of 43 NIH participants with valid task language dominance and at least one valid resting-state fMRI run.

In several participants with drug-resistant epilepsy, regions typically associated with the language network exhibited connectivity patterns characteristic of the default mode network, suggesting network reorganization. To capture this variability, we computed a resting-state laterality index (LI) based on the network topography of both the Language A and Language/ Default B networks:

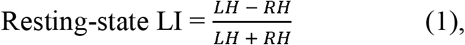

where LH and RH represent the number of vertices in the left and right hemispheres, respectively, assigned to the individual-specific Language A and Language/ Default B networks. Resting-state LI values for participants with left, bilateral, or right task-based language dominance were compared using one-way ANOVA, with p<0.05 considered statistically significant. Finally, receiver operating curves and area-under-the-curve statistics were computed to evaluate the predictive power of resting-state LI for each language dominance group.

## Results

### Participants and Datasets

We analyzed 131 participants, including 40 healthy participants from the Human Connectome Project (HCP), 65 participants with drug-resistant epilepsy from the National Institute of Health (NIH) epilepsy center, and 26 participants with drug-resistant epilepsy from the University of Iowa (esfmri dataset). HCP participants had a mean age of 29.0 ± 4.0 years and 16 (40.0%) were male. NIH participants had a mean age of 31.9 ± 11.2 years and 33 (50.8%) were male. 42 (64.6%) participants had temporal lobe epilepsy and 23 (35.4%) had extra-temporal lobe epilepsy (Table S1). Esfmri participants (Thompson et al. 2020) had a mean age of 35.8 ± 11.5 years, and 17 (65.4%) were male. 19 (73.1%) participants had temporal lobe epilepsy, 6 (23.1%) had extra-temporal lobe epilepsy, and 1 (3.8%) had undetermined seizure onset zone (Table S2). Further details regarding the data collection and pre-processing are provided in the Methods.

### Model Training and Testing

We analyzed 40 HCP and 48 NIH participants that had two or more resting-state fMRI runs to train and test MS-HBM. Participants used for training MS-HBM models were kept separate from participants used for testing the quality of individual-networks estimated by MS-HBM. Of the 48 NIH participants, 34 (70.8%) were used for the training dataset, and 14 (29.2%) were used for testing dataset (Figure 1A). To train MS-HBM models, we first estimated group-average networks of the 40 HCP participants and 34 NIH participants (Yeo et al. 2011). We also compared our results to group-average networks derived from an independent dataset of 15 intensively-sampled healthy participants from Du et al (Du et al. 2024).

Group-average networks in drug-resistant epilepsy were categorized into 9 canonical groups (Visual, Somatomotor, Auditory, Limbic, Control, Dorsal Attention, Salience/Ventral Attention, Language, and Default), which were broadly consistent with prior literature (Yeo et al. 2011, Buckner et al. 2013, Kong et al. 2019, Du et al. 2024). Group-average networks in healthy participants were more spatially distributed than those in drug-resistant epilepsy (Figure 1B). For example, association networks such as the control network were subdivided into three networks in HCP data (Kong et al. 2019), but were represented by a single, less distributed network in drug-resistant epilepsy. Instead, two spatially focal limbic networks emerged in orbitofrontal and parahippocampal regions. These differences were not explained by motion as the average relative root mean square framewise displacement was 0.059 ±0.023 vs. 0.076 ± 0.015 and the average voxel-wise differentiated signal variance (DVARS) was 23.0 ± 4.6 vs. 58.4 ± 6.8 for the NIH dataset compared to the HCP dataset respectively.

Next, we computed MS-HBM parameters using data from either (1) the 40 HCP participants and the HCP group-average (HCP MS-HBM); (2) the 40 HCP participants and the Du group-average (Du MS-HBM); or (3) the 34 NIH participants and the NIH group-average (NIH MS-HBM). After training MS-HBM, each model was then used to estimate individual-specific networks in the NIH participants with drug-resistant epilepsy. Using NIH MS-HBM, individual-specific networks captured inter-individual variability while preserving canonical topographical features of healthy participants. Notably, some participants showed altered language network organization: inferior frontal language regions exhibited stronger connectivity with middle temporal and temporopolar areas typically associated with the default mode network in healthy individuals (Figure 2A vs. 2B). These differences in language network connectivity were visually confired using identical seeds placed in the inferior frontal and lateral temporal regions across participants (Figure 2C). Quantitatively, inter-subject variability was highest in the association networks such as the salience/ventral attention, dorsal attention, control, and language networks (DICE similarity coefficient: 0.35– 0.50), as compared to the somatomotor and visual networks (DICE similarity coefficient: 0.66 – 0.71; Figure 2D).

**Figure 2:**
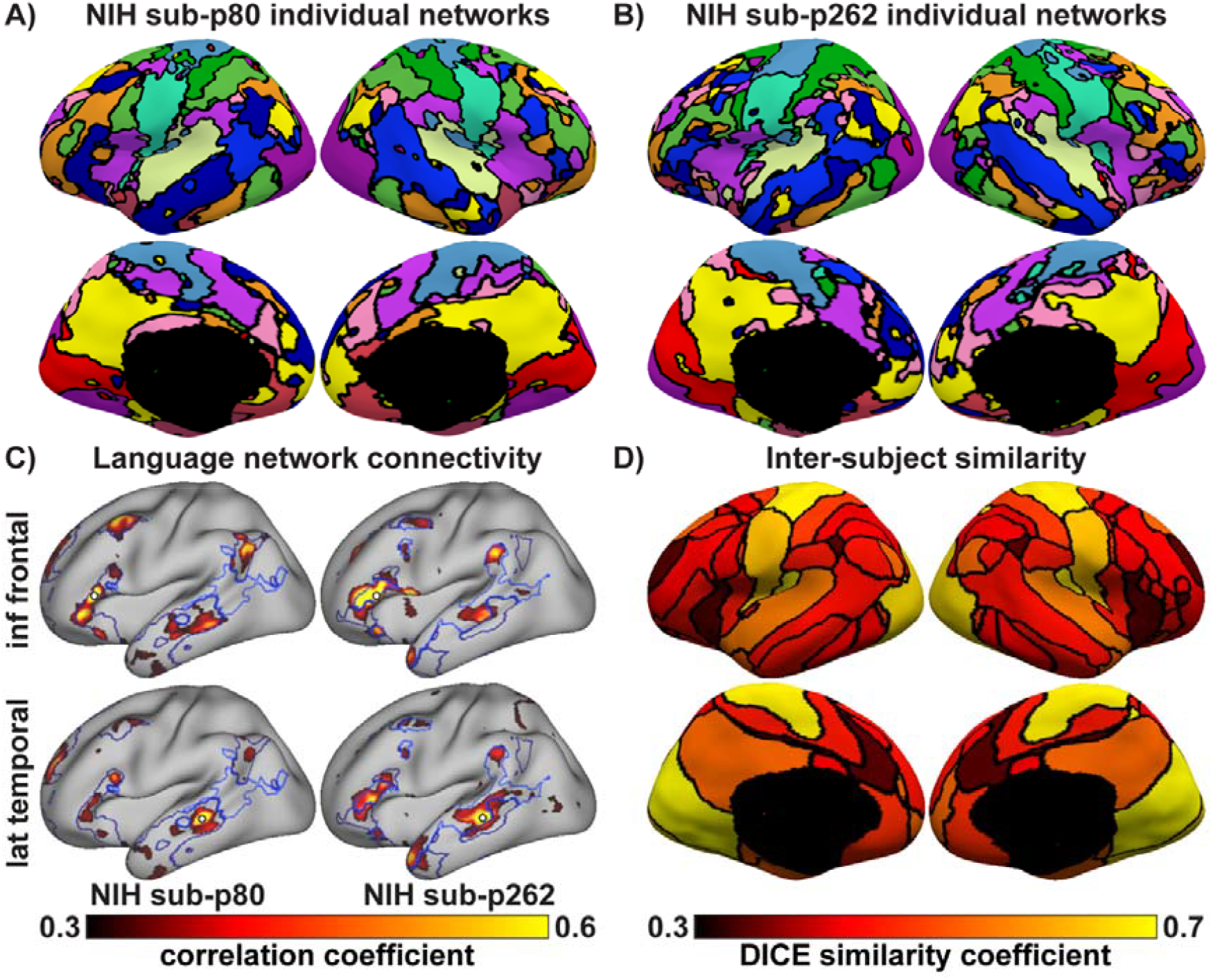
Individual-specific resting-state cortical networks in drug-resistant epilepsy. (A-B) Individual-specific networks of two representative NIH participants estimated using NIH MS-HBM. (C) Language network boundaries of both participants showing functional connectivity from identical seeds placed in the inferior frontal and lateral temporal region to different language networks in the two subjects. (D) Dice similarity coefficient for the 15-networks across all 57 NIH participants. Higher dice coefficients (yellow) indicate greater inter-subject topographic similarity, while lower dice coefficients (dark red) indicate higher variability. Inter-subject variability in drug-resistant epilepsy is higher in association than sensory networks.

Finally, in the held-out set of 14 NIH participants, we compared the quality of individual-specific networks estimated from the three MS-HBM models and group-average networks from the HCP data, Du atlas, or NIH data using an established metric, resting-state connectional homogeneity(Gordon et al. 2016, Gordon et al. 2017, Schaefer et al. 2018, Kong et al. 2019). To ensure that runs used for estimating individual-specific networks were independent from runs used for computing evaluation metrics, we used a leave-one-run-out cross-validation method. For each participant with n valid runs, individual-specific networks were estimated using n-1 runs, and evaluation metrics were computed on the left-out run. This was repeated for all runs, and an average result was obtained for each participant. We observed that individual-specific networks estimated using NIH MS-HBM performed significantly better than all group-average networks (HCP group: p<0.001; Du group: p<0.001; NIH group: p=0.003) and MS-HBM trained on healthy participants (HCP MS-HBM: p<0.001; Du MS-HBM: p<0.001), even after correcting for multiple comparisons (Figure 3A).

**Figure 3:**
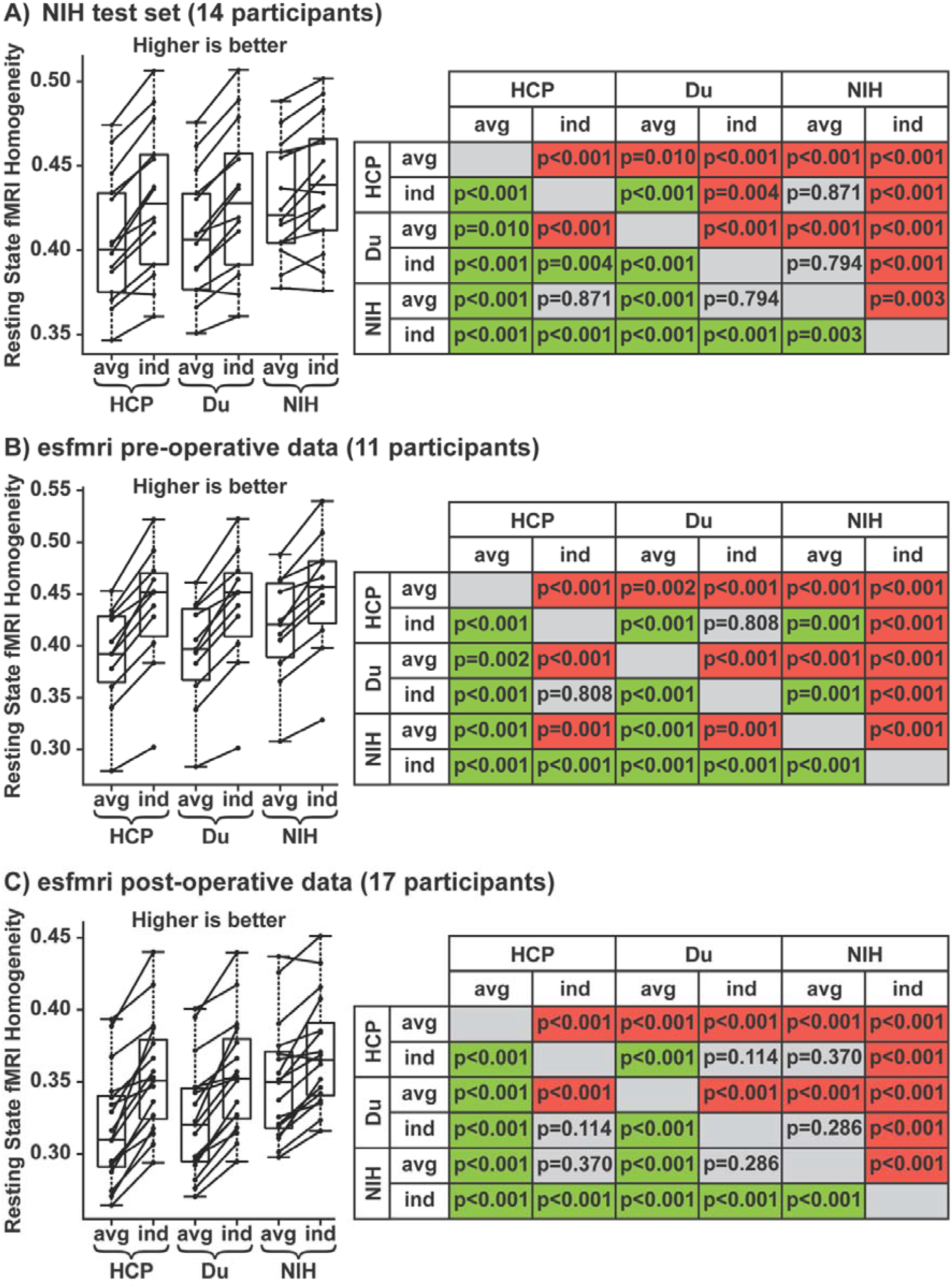
Resting-state homogeneity of group-average and individual-specific networks. Individual-specific networks estimated using NIH MS-HBM outperformed group-average networks or MS-HBM trained on healthy participants. Results were consistent across the (A) NIH test set, (B) esfmri pre-operative data, and (C) esfmri post-operative data. Six different networks were evaluated: (1) group-average and (2) individual-specific networks estimated from the HCP data; (3) group-average and (4) individual-specific networks estimated from the Du atlas; (5) group-average and (6) individual-specific networks estimated from the NIH data. Left: boxplots of median resting-state homogeneity. Right: t-test p-values comparing pairs of approaches. Green indicates that the approach on the row performed significantly better than the column, red indicates worse, and grey indicates non-significant differences, after correcting for multiple comparisons using false discovery rate of q < 0.05.

### Validation in an independent dataset

MS-HBM trained on healthy participants might perform worse than NIH MS-HBM because of site and scanner differences. Therefore, we tested if NIH MS-HBM was generalizable to an independent dataset of drug-resistant epilepsy (Thompson et al. 2020). In the esfmri dataset, there were 11 participants that had at least two pre-operative runs and 17 participants that had at least two post-operative runs (Figure 1A). We observed that individual-specific networks estimated using NIH MS-HBM performed significantly better in resting-state homogeneity than all group-average networks and MS-HBM trained on healthy participants (p<0.001 for all comparisons using either the pre-operative or post-operative data), even after correcting for multiple comparisons (Figure 3B and 3C).

In the esfmri dataset, post-operative fMRI was acquired with concurrent intracranial electrical stimulation delivered through stereoencephalography electrodes which was interleaved with echo planar imaging volume acquisition using alternating blocks of 30 seconds of stimulation or no stimulation (Thompson et al. 2020). Thus, we used a second established metric, task (electrical-stimulation) inhomogeneity, to compare the quality of the individual-specific or group-average networks. Cortical evoked activity during electrical stimulation was computed using generalized linear models (Oya et al. 2017) and electrical-stimulation inhomogeneity was computed using the average standard deviation of cortical activation z-scores within each network while accounting for network size (Gordon et al. 2016, Schaefer et al. 2018, Kong et al. 2019). A higher standard deviation indicated higher task inhomogeneity.

We observed that individual-specific networks estimated using NIH MS-HBM performed significantly better than all group-average networks for electrical-stimulation inhomogeneity (HCP group: p=0.004; Du group: p=0.005; NIH group: p=0.032), even after correcting for multiple comparisons (Figure 4A). During concurrent intracranial electrical stimulation and fMRI scans, cortical evoked activity aligned better to individual-specific network boundaries than group-average network boundaries (Figure 4B). Taken together, these results suggest that NIH MS-HBM estimate high-quality, generalizable individual-specific networks that captured the functional organization of the cortex better than group-average networks or MS-HBM trained using healthy participants.

**Figure 4:**
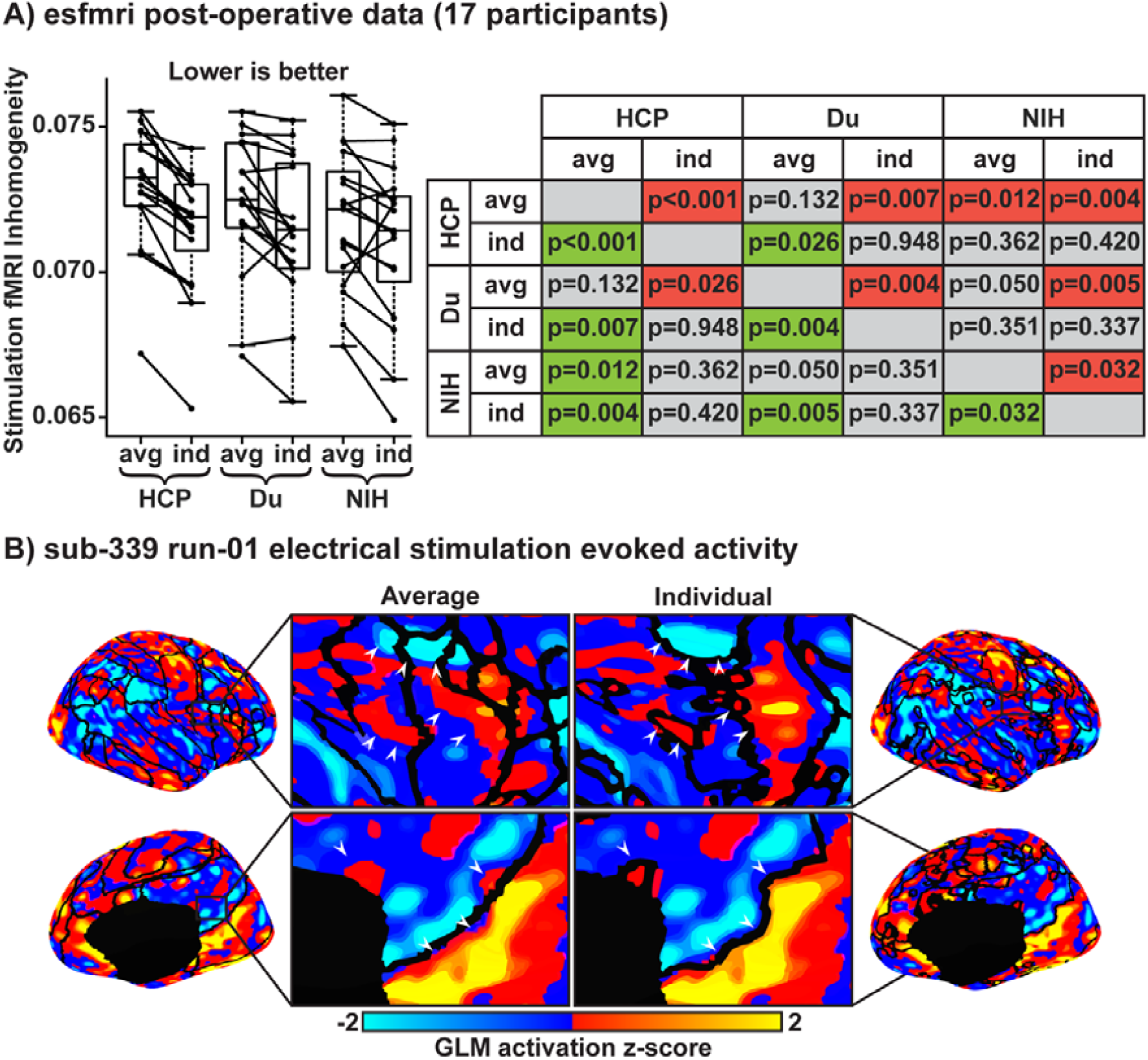
Electrical-stimulation inhomogeneity and evoked cortical activation map. Individual-specific networks estimated using NIH MS-HBM performed the best in electrical-stimulation inhomogeneity and network boundaries aligned better to electrical-stimulation evoked activity compared to group-average networks. (A) Left shows boxplots of median electrical stimulation inhomogeneity across the six networks: (1) group-average and (2) individual-specific networks estimated from the HCP data; (3) group-average and (4) individual-specific networks estimated from the Du atlas; (5) group-average and (6) individual-specific networks estimated from the NIH data. Right: t-test p-values comparing pairs of approaches. Green indicates that the approach on the row performed significantly better than the column, red indicates worse, and grey indicates non-significant differences, after correcting for multiple comparisons using false discovery rate of q < 0.05. (B) Generalised linear model of electrical-stimulation during fMRI showing z-score normalised beta-coefficients overlayed with the NIH group-average network boundaries on the left and the participant’s individual-specific network boundaries on the right. White arrowheads show evoked activity that aligned better with individual-specific networks than group-average networks.

**Figure 5:**
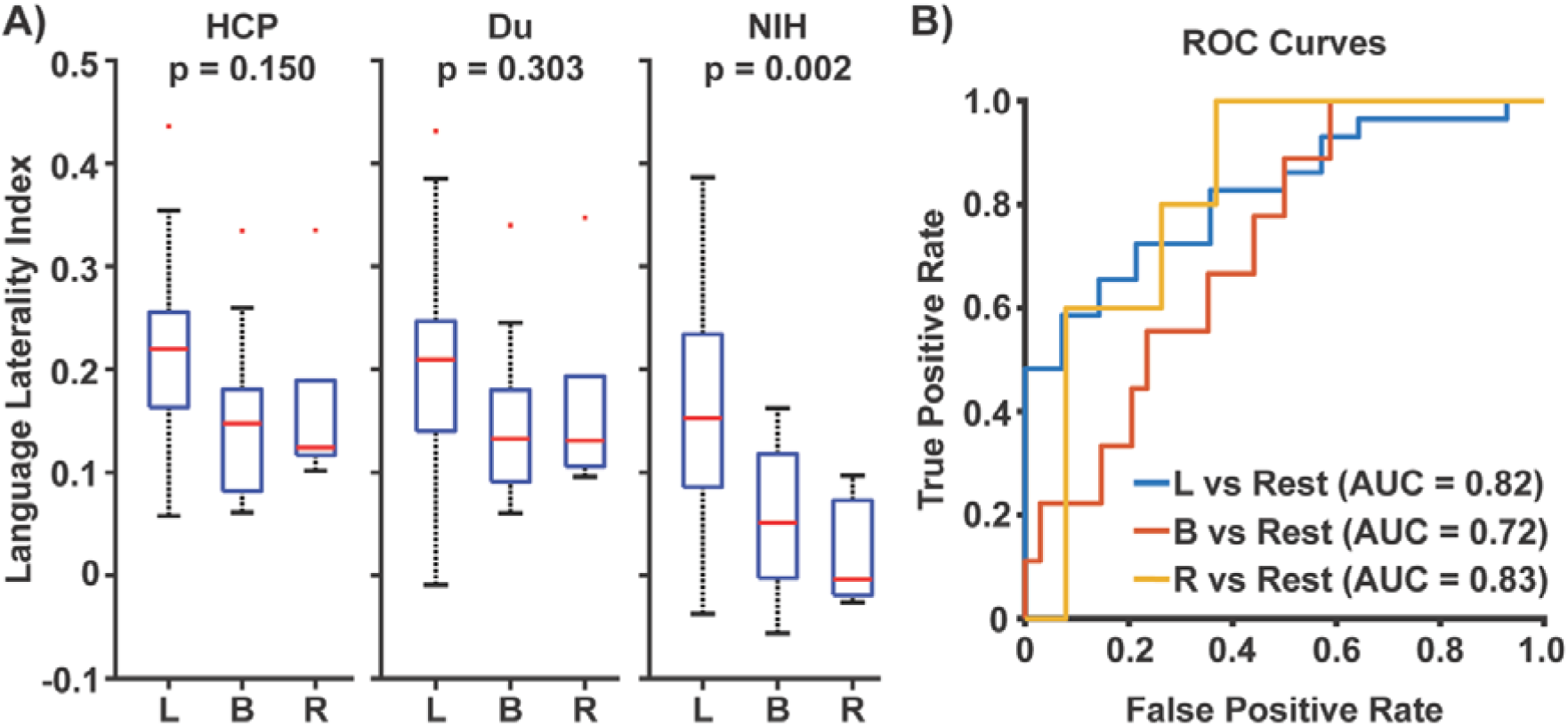
Individual-specific language network topography predicts task-based language dominance. (A) Boxplots of language laterality index for participants with left, bilateral, or right language dominance derived from task fMRI, comparing individual-specific networks estimated from HCP MS-HBM, Du MS-HBM, or NIH MS-HBM. (B) Receiver operating curves and area-under-the-curve statistic for predicting language dominance using individual-specific networks estimated by NIH MS-HBM.

### Prediction of task-based language dominance

Having shown that individual-specific networks captured the functional organization of the cortex, we next tested if language network topography using resting-state laterality index (LI) was able to predict task-based language dominance. Resting-state LI differed significantly across left (mean LI = 0.165 ± 0.106), bilateral (mean LI = 0.056 ± 0.074), and right (mean LI = 0.023 ± 0.055) task-based language dominance (p = 0.002). Receiver operating curve analyses showed area-under-the-curve statistics of 0.82 for left, 0.72 for bilateral, and 0.83 for right language dominance versus other participants. Conversely, individual-specific networks estimated using MS-HBM trained on healthy participants were not able to differentiate between task-based language dominance (HCP MS-HBM: p = 0.150; Du MS-HBM: p = 0.303).

## Discussion

In this study, we demonstrate that MS-HBM can reliably estimate individual-specific networks in drug-resistant epilepsy using only 6 to 24 minutes of resting-state fMRI data, despite altered structural and functional connectivity. Moreover, NIH MS-HBM captured the organization of an individuals’ cerebral cortex better than group-average networks and generalized to an external dataset of drug-resistant epilepsy despite site, scanner, and pre-processing differences. Finally, we showed that individual-specific language network topography predicts task-based language dominance well in individual patients with drug-resistant epilepsy.

Existing evidence indicates that focal epilepsy is associated with reduced network integration and increased segregation, changes that may underlie the cognitive and behavioral comorbidities commonly observed in epilepsy (van Diessen et al. 2014, Courtiol et al. 2020). Although approaches such as coordinate-based network mapping have successfully delineated an idiopathic generalized epilepsy network (Ji et al. 2025), they are limited in their ability to characterize epilepsy-related reorganization of intrinsic resting-state networks at the individual level. As a result, cortical network organization in drug-resistant epilepsy, particularly alterations to canonical resting-state networks, remains incompletely characterized (Caciagli et al. 2014). Consistent with these limitations, prior resting-state fMRI approaches were unable to reliably estimate individual-specific cortical organization of the language networks and predict task-based language dominance in drug-resistant epilepsy (Doucet et al. 2015, Rolinski et al. 2020, Phillips et al. 2021, Pur et al. 2021). This gap likely reflects both the limited availability of high-quality resting-state fMRI data in clinical populations and the substantial heterogeneity of pathology in drug-resistant epilepsy (Sisodiya et al. 2020).

By leveraging MS-HBM, we begin to reveal the functional organization of focal drug-resistant epilepsy, with a particular focus on language networks. While cortical networks in drug-resistant epilepsy broadly resemble canonical resting-state networks observed in healthy participants, their spatial topography is more focal (Liao et al. 2010, Bernhardt et al. 2011), and individual-level variations emerge that are not typically seen in healthy brains. These differences cannot be explained by motion. Notably, inferior frontal language regions in some participants exhibit stronger connectivity with areas typically associated with the default mode network in healthy individuals, potentially reflecting the long-term effects of chronic epileptogenic activity on functional connectivity (van Diessen et al. 2013, Englot et al. 2016). By capturing these inter-individual variations in language network topography, MS-HBM enables reliable prediction of task-based language dominance. In this context, precision functional mapping provides a powerful approach to elucidate how chronic epileptogenic activity reorganizes resting-state cortical networks and influences cognitive function (Robert et al. 2024).

Future studies should validate the resting-state laterality index derived from MS-HBM against gold-standard measures, such as the intracarotid amobarbital test (Binder et al. 1996) or cortical electrical stimulation (Austermuehle et al. 2017). While this study focused on language networks and their role in predicting language dominance, these results more broadly demonstrate that MS-HBM can reliably estimate individual-specific networks in patients with neurological disorders, even with limited resting-state fMRI data. This approach could be extended to predict other cognitive comorbidities (Englot et al. 2016, Zhang et al. 2020, Ren et al. 2022, Royer et al. 2023, Ankeeta et al. 2025) and post-operative functional outcomes (Bettus et al. 2010, Boerwinkle et al. 2017, Boerwinkle et al. 2019, Akbarian et al. 2024, Wang et al. 2024) in patients with drug-resistant epilepsy. Together, these findings suggest that MS-HBM may be applied to other clinical populations and neurological pathologies, offering a practical, non-invasive tool for presurgical evaluation and potentially guiding individualized clinical decision-making (Boerwinkle et al. 2020).

Several limitations of our study should be acknowledged. The esfmri dataset provided a unique opportunity to validate individual-specific networks using evoked activity during concurrent intracranial electrical stimulation (Thompson et al. 2020). However, this dataset did not collect true post-operative resting-state fMRI, and we concatenated blocks of fMRI data when no stimulation was applied to simulate resting-state scans (Pedersen et al. 2021). Comparisons of functional connectivity within the same participants across pre- and post-operative data revealed differences that may reflect (1) the effects of neurosurgery, (2) signal distortion due to implanted electrodes, or (3) scanner-related differences. Despite these factors, NIH MS-HBM consistently produced high-quality individual-specific networks across both pre- and post-operative data in the esfmri dataset. Finally, we studied participants with temporal lobe and extra-temporal lobe epilepsy undergoing presurgical evaluation, and future studies are required to validate these networks in other types of drug-resistant epilepsy (e.g. idiopathic generalized epilepsy).

## Conclusion

In summary, we present a method for estimating reliable, high-quality individual-specific cortical networks in drug-resistant epilepsy and showed that individual-specific language network topography predicts task-based language dominance. Our model is publicly available (github link), which may be used to estimate individual-specific cortical networks and predict language dominance in a new individual with drug-resistant epilepsy given at least 6 minutes of single-session resting-state fMRI data.

## Data Availability

Data from the NIH produced in the present study are available upon reasonable request to the authors. Data from the esfmri dataset and human connectome project may be obtained online.

https://openneuro.org/datasets/ds002799/versions/1.0.4

https://www.humanconnectome.org/

## Data Availability

Deidentified data from the National Institutes of Health will be made available upon reasonable request to. All other data used are open source and available at their respective sites.

## Acknowledgements

This study was supported in part by NIH-DIR ZIA NS009431-05 from the Division of Intramural Research (DIR) at the National Institute of Neurological Disorders and Stroke, National Institutes of Health, Bethesda, MD, USA.

## Disclosure of Conflicts of Interest

None of the authors has any conflict of interest to disclose.

## Notes

### Competing Interest Statement

The authors have declared no competing interest.

### Author Declarations

Ethics committee of the NIH Combined Neurosciences Institutional Review Board and University of Iowa Institutional Review Board gave ethical approval for this work.

### Summary of Updates

We have provided further details on the methods and expanded on the introduction and discussion in this version of the manuscript.

