## Supplemental Methods for "Individual-specific resting-state networks predict language dominance in drug-resistant epilepsy"

### Supplementary Methods

#### Participants and Datasets

The HCP dataset consisted of 40 participants from the Human Connectome Project (HCP) S900 release. The mean age was 29.0 ± 4.0 years, there were 16 (40.0%) male participants, and 12 (30.0%) participants self-reported as non-White and/or Hispanic race. Each subject had one 0.7mm isotropic T1 structural scan and four runs of 2mm isotropic resting-state fMRI with a TR of 0.72s and lasting 14min 33s each. All imaging data were collected on a custom-made Siemens 3 T Skyra scanner using a multiband sequence. Details of the data collection may be found elsewhere (Van Essen et al. 2012, Smith et al. 2013).

The NIH dataset consisted of 65 participants with drug-resistant epilepsy undergoing presurgical evaluation at the NIH epilepsy center (Supplementary Table 1). The mean age of participants was 31.9 ± 11.2 years, and there were 33 (50.8%) male participants. There were 42 (64.6%) participants with temporal lobe epilepsy, and 23 (35.4%) participants with extra-temporal lobe epilepsy. All participants gave informed written consent under a research protocol approved by the NIH Combined Neurosciences Institutional Review Board. Each participant had one 0.8 x 0.75 x 0.75mm T1 and T2 structural scans, up to four runs of 3mm isotropic resting-state fMRI with a TR of 2.5s and lasting 6min 25s each, and a 3mm isotropic auditory description decision language task fMRI run with a TR of 2.0s and lasting 5min. All imaging data were collected on a 3.0 Tesla scanner at the NIH Nuclear Magnetic Resonance Center using a multiband sequence on either a Siemens Skyra scanner or a GE Discovery scanner. Details of the data collection may be found elsewhere (Rolinski et al. 2020).

The esfmri dataset (Thompson et al. 2020) consisted of 26 participants with drug-resistant epilepsy undergoing presurgical evaluation at the University of Iowa (Supplementary Table 2). The mean age of participants was 35.8 ± 11.5 years, and there were 17 (65.4%) male participants. There were 19 (73.1%) participants with temporal lobe epilepsy, 6 (23.1%) participants with extra-temporal lobe epilepsy, and 1 (3.8%) participant where the seizure onset zone was not determined. Intracranial electrodes were implanted to localize the epileptogenic zone, and were determined by clinical criteria. All participants gave informed written consent under a research protocol approved by the University of Iowa Institutional Review Board (Thompson et al. 2020). Each participant had one 1.0 x 1.0 x 0.8mm T1 structural scan, up to five runs of 3.4 x 3.4 x 4.0mm pre-operative resting state fMRI with a TR of 2.26s lasting 4.8min each, and up to eleven runs of 3mm isotropic electrical-stimulation fMRI with a TR of 3.0s lasting approximately 10min each. For each post-operative fMRI scan, bipolar intracranial electrical stimulation using depth electrodes was interleaved between echo planar imaging volume acquisition during a 100ms period where there was no scanner radiofrequency or gradient switching (Thompson et al. 2020). Stimulation was blocked with alternating durations of approximately 30 seconds of STIM-ON and STIM-OFF each. No cognitive task was given to the participant, and no behavioural effects were evoked during the stimulation period. Details of the data collection may be found elsewhere (Oya et al. 2017, Thompson et al. 2020).

#### Pre-processing of Imaging Data

The HCP dataset was released with data already pre-processed (smoothed using a 2mm full-width half-maximum kernel, denoised with ICA-FIX and aligned with MSMAll) and were detailed elsewhere (Kong et al. 2019, Du et al. 2024). The preprocessed fMRI data was projected onto the FreeSurfer fsaverage6 surface space. Additional nuisance regression, censoring, and spatial smoothing was computed using the surface-based pipeline of Yeo et al (Kong et al. 2019). After pre-processing, there were 40 HCP participants with 4 runs each.

Pre-processing of the NIH dataset followed the surface-based pipeline of Yeo et al (Yeo et al. 2011, Holmes et al. 2015). Structural data were processed using recon-all (FreeSurfer 7.1.1) (Dale et al. 1999). Resting-state fMRI data were processed with the following: (i) removal of first 4 frames, (ii) slice time correction using FSL (Jenkinson et al. 2002), (iii) motion correction and outlier detection using rigid body translation and rotation (Jenkinson et al. 2002), (iv) susceptibility-induced spatial distortion correction (Andersson et al. 2003), (v) multi-echo denoising (DuPre et al. 2021), (vi) boundary-based registration (Greve et al. 2009), (vii) regression of the global, white matter, ventricular signals, 6 motion parameters, and their temporal derivatives, (viii) censored frames were interpolated with the Lomb-Scargle periodogram (Power et al. 2014), (ix) bandpass filtering of 0.009Hz – 0.08Hz, and lastly, (x) the data was projected onto FreeSurfer fsaverage6 surface space and smoothed using a 6mm full-width half-maximum kernel. Outlier detection using framewise displacement (FD) and voxel-wise differentiated signal variance (DVARS) were computed using fsl_motion_outliers (Smith et al. 2004). Frames that exceeded a threshold of 0.2mm FD or 50 DVARS as well as one frame before and two frames after were censored. Segments of data lasting less than five contiguous frames were also censored (Gordon et al. 2016). After pre-processing, 8 participants did not have more than one valid resting-state run, 9 participants had one run, 23 participants had two runs, 24 participants had three runs, and one participant has four runs. Of the 48 NIH participants with two or more runs, 34 (70.8%) participants were used for the training set, and 14 (29.2%) were used for the test set.

The esfmri dataset was released with data pre-processed using fMRIPrep 1.5.1rc1 (Esteban et al. 2019) (based on Nipype 1.2.0 (Gorgolewski et al. 2011)) and was described in detail elsewhere (Thompson et al. 2020). Frames that exceeded a threshold of 0.2mm FD or 1.2 standardized derivative of the root-mean-square variance were censored. Runs that had more than 50% motion outliers were discarded. Linear regression was used to regress out 6 motion parameters, the whole brain, white matter, and cerebrospinal fluid regressors and their derivatives. The fMRI time-series were then resampled to the FreeSurfer fsaverage6 space and spatially smoothed using a 6mm full-width half-maximum kernel (FreeSurfer 5.3.0) (Dale et al. 1999). After pre-processing, there were 11 participants that had at least two valid pre-operative runs and 17 participants that had at least two valid post-operative runs.

#### Derivation of group-average networks and training of MS-HBM for estimating individual-specific networks

Our approach has been described previously (Yeo et al. 2011, Kong et al. 2019). Briefly, the pre-processed fMRI data were projected onto fsaverage6 surface meshes that consisted of 40,962 vertices per hemisphere. The connectivity profile of a cortical region (vertex) was defined to be its functional coupling to 1175 regions of interest (ROIs), which consisted of single vertices uniformly distributed across the fsaverage6 surface meshes. For each run, the Pearson’s correlation between the fMRI time series at each spatial location (81,924 vertices) and the 1175 ROIs were computed. The 81,924 x 1175 correlation matrix was then binarized by keeping the top 10% of correlations. To obtain group-average networks, each vertex’s connectivity profiles were averaged across all resting-state fMRI runs of all participants. The averaged connectivity profiles were then clustered using a mixture of von Mises-Fisher distributions to obtain the 15-network group-average for each dataset (Lashkari et al. 2010, Yeo et al. 2011). In addition to computing group-average networks for the 40 HCP participants and 34 NIH participants, we also compared our results to group-average networks derived from an independent dataset of 15 intensively-sampled healthy participants (Du et al. 2024).

We trained MS-HBM using either 40 participants from the Human Connectome Project (HCP dataset), or 34 participants with drug-resistant epilepsy from the National Institutes of Health (NIH dataset). In addition, we used a group-average atlas of 15 densely sampled healthy participants (Du et al. 2024) to train MS-HBM on the 40 participants from the HCP dataset. Model parameters estimated included the inter-subject functional connectivity variability, intra-subject functional connectivity variability, spatial smoothness prior, and the inter-subject spatial variability prior (Kong et al. 2019). Using each of the three MS-HBM models, individual-specific networks were estimated for each participant with drug-resistant epilepsy. For all individual-specific networks estimated, the weight of the group spatial prior was taken to be 80, and the weight of the smoothness prior was taken to be 10 (Kong et al. 2019). Individual-specific networks were visually confirmed using model-free seed-based functional connectivity. Inter-subject similarity in network topography across the 15-networks was quantified using the Dice similarity coefficient across all 57 NIH participants with at least one valid run.

#### Generalized linear models of intracranial electrical stimulation during fMRI scans

Generalized linear models were used to quantify the effects of electrical stimulation on cortical activity. A boxcar function of the stimulus duration (varies from 50-90 ms) for each post-operative run was computed (Thompson et al. 2020). The hemodynamic response function was then modelled by convolving the boxcar function with an optimised set of three gamma basis functions generated using FSL Linear Optimal Basis Sets (Woolrich et al. 2004) to account for any non-canonical haemodynamic response function shapes caused by the stimulation (parameters used were m1 = 0-2s; m2 = 3-6s; m3 = 3-8s; m4 = 3-8s; c = 0-0.3). The amplitude of all basis functions were normalized to peak values of 1 by dividing by the maximum amplitude before computing the haemodynamic response function.

The generalized linear models were computed using freesurfer’s mri_glmfit on the fsaverage6 data and corrected at the vertex-level for the lag one autoregression correlation matrix averaged across all valid runs. The vertex-level unthresholded beta-coefficients of the contrast consisting of all three haemodynamic response function shapes were then Z-score normalised by subtracting the mean and dividing by the standard deviation across all beta-coefficients of both hemispheres.
