## Supplemental Table 1 for "Individual-specific resting-state networks predict language dominance in drug-resistant epilepsy"

| **No.** | **Age** | **Sex** | **Hand** | **Seizure Onset Zone** | **pre-op MRI** | **pre-op Runs** | **post-op MRI** | **post-op Runs** |
| --- | --- | --- | --- | --- | --- | --- | --- | --- |
| 1 | 46-50 | F | L | Left mesial temporal lobe, Left frontal lobe | Siemens Trio | 0 | Siemens Trio | 5 |
| 2 | 31-35 | M | R | Right anterior frontal lobe | Siemens Trio | 0 | Siemens Trio | 2 |
| 3 | 46-50 | F | R | Left mesial temporal lobe | Siemens Trio | 0 | Siemens Skyra | 1 |
| 4 | 31-35 | F | R | Right mesial temporal lobe, Left mesial temporal encephalomalacia | Siemens Trio | 1 | Siemens Skyra | 4 |
| 5 | 26-30 | M | R | Left insula | Siemens Trio | 1 | Siemens Skyra | 1 |
| 6 | 26-30 | F | R | Bilateral mesial temporal lobe | Siemens Trio | 5 | Siemens Skyra | 0 |
| 7 | 31-35 | F | R | Right mesial temporal lobe | Siemens Trio | 4 | Siemens Skyra | 7 |
| 8 | 46-50 | F | R | Right hippocampus | Siemens Trio | 0 | Siemens Skyra | 0 |
| 9 | 41-45 | M | L | Left occipital lobe | Siemens Trio | 5 | Siemens Skyra | 4 |
| 10 | 31-35 | M | R | Left mesial temporal lobe | Siemens Trio | 1 | Siemens Skyra | 5 |
| 11 | 36-40 | M | L | Right temporal pole, Left temporal base | Siemens Trio | 5 | Siemens Skyra | 10 |
| 12 | 31-35 | M | R | Bilateral mesial temporal lobe | Siemens Trio | 3 | Siemens Skyra | 2 |
| 13 | 41-45 | M | R | Not determined | None | - | Siemens Skyra | 6 |
| 14 | 31-35 | M | B | Left frontal cystic mass | GE Discovery | 2 | Siemens Skyra | 0 |
| 15 | 36-40 | M | R | Left mesial temporal lobe | None | - | Siemens Skyra | 1 |
| 16 | 26-30 | M | R | Right mesial temporal lobe | None | - | Siemens Skyra | 8 |
| 17 | 31-35 | M | R | Left temporal pole | GE Discovery | 2 | Siemens Skyra | 0 |
| 18 | 46-50 | F | R | Right mesial temporal lobe | GE Discovery | 3 | Siemens Skyra | 2 |
| 19 | 36-40 | M | R | Right mesial temporal lobe, Right frontal pole | GE Discovery | 2 | Siemens Skyra | 0 |
| 20 | 21-25 | M | R | Right amygdala | None | - | Siemens Skyra | 7 |
| 21 | 11-15 | M | R | Left superior frontal gyrus cavernoma | None | - | Siemens Skyra | 3 |
| 22 | 21-25 | F | R | Right mesial temporal lobe, Possible right frontal base | GE Discovery | 3 | Siemens Skyra | 6 |
| 23 | 56-60 | M | R | Left mesial temporal lobe | GE Discovery | 1 | Siemens Skyra | 2 |
| 24 | 56-60 | F | R | Left mesial temporal lobe | None | - | Siemens Skyra | 0 |
| 25 | 16-20 | M | R | Left frontal encephalomalacia | GE Discovery | 4 | Siemens Skyra | 3 |
| 26 | 21-25 | M | L | Right mesial temporal lobe | GE Discovery | 5 | Siemens Skyra | 2 |

**Supplementary Table 2**: Demographic, MRI, and epilepsy characteristics of participants from the esfmri dataset.
