## Supplemental Table 2 for "Individual-specific resting-state networks predict language dominance in drug-resistant epilepsy"

| **No.** | **Age** | **Sex** | **Hand** | **Seizure Onset Zone** | **Task Laterality** | **MRI** | **Valid Runs** |
| --- | --- | --- | --- | --- | --- | --- | --- |
| 1 | 26-30 | F | R | Right Mesial Temporal Sclerosis | L | Siemens Skyra | 2 |
| 2 | 20-25 | F | R | Right Parietal | U | GE Discovery MR750 | 2 |
| 3 | 36-40 | M | R | Left > Right Temporal | B | GE Discovery MR750 | 1 |
| 4 | 30-35 | M | R | Right > Left Medial Temporal | L | Siemens Skyra | 2 |
| 5 | 20-25 | F | R | Left Mesial Temporal Sclerosis | U | GE Discovery MR750 | 1 |
| 6 | 36-40 | M | R | Right Mesial Temporal Sclerosis | L | Siemens Skyra | 0 |
| 7 | 30-35 | F | R | Right Temporoparietal | L | GE Discovery MR750 | 2 |
| 8 | 26-30 | F | R | Right Temporal | B | GE Discovery MR750 | 2 |
| 9 | 30-35 | F | R | Right Mesial Temporal Sclerosis | L | Siemens Skyra | 3 |
| 10 | 20-25 | M | L | Left Mesial Temporal Sclerosis | L | GE Discovery MR750 | 2 |
| 11 | 36-40 | F | R | Left Temporal | R | GE Discovery MR750 | 2 |
| 12 | 56-60 | F | R | Right Frontal | L | GE Discovery MR750 | 2 |
| 13 | 16-20 | M | R | Right Frontal | L | GE Discovery MR750 | 1 |
| 14 | 36-40 | F | R | Left Frontal/ Insula | U | GE Discovery MR750 | 1 |
| 15 | 20-25 | M | R | Right Frontal | U | GE Discovery MR750 | 2 |
| 16 | 30-35 | M | R | Left Temporal Periventricular Nodular Heterotopia | B | Siemens Skyra | 3 |
| 17 | 36-40 | M | L | Left > Right Frontal | U | GE Discovery MR750 | 1 |
| 18 | 36-40 | F | L | Left Mesial Temporal Sclerosis | R | GE Discovery MR750 | 2 |
| 19 | 20-25 | M | R | Left Frontal/ Insula | L | GE Discovery MR750 | 2 |
| 20 | 36-40 | M | R | Right Frontal | U | GE Discovery MR750 | 2 |
| 21 | 30-35 | F | R | Left Mesial Temporal Sclerosis | L | Siemens Skyra | 1 |
| 22 | 16-20 | F | R | Left Frontotemporal | R | GE Discovery MR750 | 2 |
| 23 | 51-55 | M | R | Left Temporal | L | Siemens Skyra | 3 |
| 24 | 26-30 | F | R | Right Temporal | R | GE Discovery MR750 | 2 |
| 25 | 30-35 | F | R | Left Temporal | L | Siemens Skyra | 2 |
| 26 | 16-20 | F | R | Right Frontal | L | GE Discovery MR750 | 2 |
| 27 | 36-40 | F | R | Left Temporal | B | GE Discovery MR750 | 0 |
| 28 | 56-60 | F | R | Left Mesial Temporal Sclerosis | L | Siemens Skyra | 3 |
| 29 | 26-30 | M | B | Bilateral Frontal | L | GE Discovery MR750 | 2 |
| 30 | 30-35 | F | R | Left > Right Temporal | L | Siemens Skyra | 3 |
| 31 | 26-30 | F | L | R Insula > Frontal | R | GE Discovery MR750 | 2 |
| 32 | 20-25 | F | B | Right Polymicrogyria, Bilateral Periventricular Nodular Heterotopia | B | GE Discovery MR750 | 2 |
| 33 | 51-55 | M | R | Left Frontal Schizencephaly | U | GE Discovery MR750 | 0 |
| 34 | 26-30 | F | R | Left Mesial Temporal Sclerosis | B | Siemens Skyra | 3 |
| 35 | 41-45 | M | R | Right Frontal | L | Siemens Skyra | 3 |
| 36 | 26-30 | F | R | Right Mesial Temporal Sclerosis | U | Siemens Skyra | 1 |
| 37 | 30-35 | M | R | Right Parietal | L | Siemens Skyra | 3 |
| 38 | 41-45 | F | L | Right Frontal | B | Siemens Skyra | 3 |
| 39 | 16-20 | F | R | Right Temporal | U | Siemens Skyra | 0 |
| 40 | 56-60 | M | R | Left Temporal | B | Siemens Skyra | 1 |
| 41 | 20-25 | M | R | Left Temporal | L | Siemens Skyra | 3 |
| 42 | 36-40 | F | R | Right Temporal | U | Siemens Skyra | 3 |
| 43 | 20-25 | M | L | Right Temporal Arteriovenous Malformation | L | Siemens Skyra | 3 |
| 44 | 16-20 | M | L | Left Parietal | B | Siemens Skyra | 3 |
| 45 | 20-25 | M | R | Left Frontal | L | Siemens Skyra | 2 |
| 46 | 26-30 | M | R | Left Temporal Low Grade Glioma | L | Siemens Skyra | 3 |
| 47 | 20-25 | F | R | Left Frontal | L | Siemens Skyra | 3 |
| 48 | 26-30 | M | R | Right Parieto-occipital | L | Siemens Skyra | 0 |
| 49 | 30-35 | M | R | Left Mesial Temporal Sclerosis | U | Siemens Skyra | 3 |
| 50 | 20-25 | M | R | Right Frontal | L | Siemens Skyra | 4 |
| 51 | 30-35 | M | L | Left Temporal | U | Siemens Skyra | 3 |
| 52 | 20-25 | F | R | Right Temporal | L | Siemens Skyra | 2 |
| 53 | 26-30 | F | R | Left Temporal | U | Siemens Skyra | 3 |
| 54 | 56-60 | M | R | Right Temporal | L | Siemens Skyra | 1 |
| 55 | 6-10 | M | R | Left Frontal | L | Siemens Skyra | 3 |
| 56 | 26-30 | M | R | Bilateral Temporal | U | Siemens Skyra | 3 |
| 57 | 36-40 | M | R | Left Temporal | U | Siemens Skyra | 3 |
| 58 | 20-25 | M | R | Right Temporal | L | Siemens Skyra | 3 |
| 59 | 51-55 | F | R | Right Temporal | L | Siemens Skyra | 3 |
| 60 | 30-35 | F | R | Right Temporal | R | Siemens Skyra | 0 |
| 61 | 20-25 | F | R | Right Parietal/ Temporal | L | Siemens Skyra | 0 |
| 62 | 26-30 | M | R | Left Temporal | U | Siemens Skyra | 3 |
| 63 | 26-30 | M | R | Right Temporal | L | Siemens Skyra | 2 |
| 64 | 51-55 | F | R | Left Temporal | L | Siemens Skyra | 0 |
| 65 | 30-35 | M | R | Left Temporal | B | Siemens Skyra | 2 |

**Supplementary Table 1**: Demographic, MRI, and epilepsy characteristics of participants from the NIH dataset.
